# Estimating the Impact of Self-Management Education, Influenza Vaccines, Nebulizers, and Spacers on Healthcare Utilization and Expenditures for Medicaid-Enrolled Children with Asthma

**DOI:** 10.1101/2020.09.28.20188466

**Authors:** Melike Yildirim, Paul Griffin, Pinar Keskinocak, Jean O’Connor, Julie L Swann

## Abstract

We quantify the effect of a set of interventions including asthma self-management education, influenza vaccination, spacers, and nebulizers on healthcare utilization and expenditures for Medicaid-enrolled children with asthma in New York and Michigan.

**Methods:** We obtained patients’ data from Medicaid Analytic eXtract files and evaluated patients with persistent asthma in 2010 and 2011. We used difference-in-difference regression to quantify the effect of the intervention on the probability of asthma-related healthcare utilization, asthma medication, and utilization costs. We estimated the average change in outcome measures from pre-intervention/intervention (2010) to post-intervention (2011) periods for the intervention group by comparing this with the average change in the control group over the same time horizon.

**Results:** All of the interventions reduced both utilization and asthma medication costs. Asthma self-management education, nebulizer, and spacer interventions reduced the probability of emergency department (20.8-1.5 %, 95%CI 19.7-21.9% vs. 0.5-2.5% respectively) and inpatient (3.5-0.8%, 95%CI 2.1-4.9% vs. 0.4-1.2%, respectively) utilizations. Influenza vaccine decreased the probability of primary care physician (6-3.5%, 95%CI 4.4-7.6% vs. 1.5-5.5%, respectively) visit. The reductions varied by state and intervention.

**Conclusions:** Promoting asthma self-management education, influenza vaccinations, nebulizers, and spacers can decrease the frequency of healthcare utilization and asthma-related expenditures while improving medication adherence.

## Introduction

Asthma is a chronic inflammatory disease of the airways that causes wheezing, breathlessness, and chest tightness (1). Asthma is one of the most common chronic illnesses, affecting 8.3% of US children in 2016 (2). The estimated annual cost of asthma treatment in the US was $81.9 billion in 2013, with an average of $3,728 per person (3). Additionally, asthma is the third leading cause of pediatric hospitalization (4), and children missed 13.8 million school days in 2013 due to asthma complications (5). Asthma severity may change over time. However, with routine follow up visits to the primary physician as well as careful adherence to medications and avoidance of asthma triggers, patients can prevent and control asthma symptoms (6). If the patient has uncontrolled asthma and the symptoms are present, it is crucial to optimize care (e.g., asthma self-management education (AS-ME)).

Improvements to asthma treatment and education can reduce the frequency of related emergency department (ED) visits, missed school days, and hospitalizations. The Center for Disease and Control (CDC) prioritized several National Asthma Education and Prevention Program (NAEPP) recommendations in 6|18 initiative, which includes access and adherence to the appropriate asthma medications and devices and promotes AS-ME for individuals whose asthma is not well controlled (7).

A nebulizer is a device to deliver liquid medication to the lungs accurately, and spacers are recommended to use with inhalers to provide a required amount of medication, especially by children who have difficulty inhaling correctly. Proper usage of medications and required dose leads to a better response (8). Two types of medications (controllers and relievers) are commonly used for asthma treatments. Relievers work on acute symptoms and provide quick relief, whereas controller medications are used regularly to prevent asthma attacks. Improving adherence to asthma medications and (9). For patients whose asthma remains uncontrolled despite medical management, AS-ME helps patients manage asthma triggers and symptoms and reduce asthma exacerbation (10). Patients with asthma have an increased risk of severe symptoms with influenza. Flu vaccines can decrease the risk of asthma attacks, which is triggered by flu infection (11). Although clinical guidelines have recommended AS-ME and prescription of a spacer with an inhaler (7), they are not widely implemented, and their net benefits are uncertain.

The objectives of this study were to: (i). predict the effects of asthma interventions (AS-ME programs, influenza vaccination, nebulizer and spacer) on healthcare utilization cost and asthma medication expenditures; (ii). evaluate the impacts of implementing the recommended asthma interventions on the likelihood of asthma-related utilization (ED visits, inpatient (IP) visits, and primary care physician (PCP) visits). Our study has several strengths. We use the difference-in-difference method, which allows us to predict the net effect of interventions while controlling for various time-dependent factors. We also analyze several types of interventions (AS-ME, influenza vaccination, nebulizer and spacer) used by children in two Medicaid programs across several performance measures. The results provide insight for programs considering such interventions.

## Methods

### Data and Study Population

We evaluated claims data from children in New York and Michigan aged 0-17 with persistent asthma in 2010 and 2011 in the Medicaid program. Patients were Medicaid-eligible and enrolled for the entire 24 months. Based on slightly modified (12) criteria defined by Healthcare Effectiveness Data and Information Set (HEDIS), we defined persistent asthma patients as those who have at least one prescription of asthma controller medication or at least one ED visit or hospitalization, or at least three outpatient visits with a primary diagnosis of asthma. We used HEDIS’s definition for controller medication (including anti-asthmatic combinations, antibody inhibitors, inhaled corticosteroids, inhaled steroid combinations, leukotriene modifiers, mast cell stabilizers, and methylxanthines) (13). We classified patients with diagnoses of asthma using the ICD-9 (493.XX) codes. We identified 59,685 and 14,358 patients, respectively, in NY and MI.

We selected New York and Michigan from a set of ten states (Texas, New York, California, Florida, North Carolina, Alabama, Ohio, Illinois, Montano, and Michigan) with a high number of claims for AS-ME in 2010. We chose MI and NY because they also had high utilization of spacers and nebulizers, and they have different characteristics for comparison purposes.

We utilized all patients’ data, asthma-related expenditures, utilizations, and interventions in 2010 and 2011 from Medicaid Analytic eXtract (MAX) files. We obtained the urban, suburban, and rural classification of each zip code from the 2010 Census data (14).

We examined four interventions: AS-ME, influenza vaccine, nebulizer, or spacer interventions in 2010. We evaluated one intervention effect in each model. We defined four treatment subpopulations, i.e., one for each intervention: i) AS-ME intervention patients - had at least one claim file with the procedure of AS-ME in 2010 with a primary diagnosis code for asthma, ii) influenza intervention patients - had a prescription of influenza vaccine in 2010, iii) spacer intervention patients - had at least one prescription for a spacer (spacer with and without a mask or holding chamber) in 2010, iv) nebulizer intervention patients - had at least one prescription for a nebulizer in 2010, and v) patients who did not receive the specified intervention in either year. For example, the control population of AS-ME consisted of the patients who did not receive AS-ME in 2010 and 2011. Similarly, each intervention population had corresponding control groups (four control populations: AS-ME, influenza vaccine, nebulizer, and spacer controls).

Patients were excluded from our analysis if they received asthma-related home visits in 2010 or 2011. We dropped intervention related utilizations and costs (e.g., visits for AS-ME) from the data.

### Dependent Variables

We focus on utilization associated with active management of asthma, such as in a primary care setting. We are also concerned with the most expensive visits, including those in the ED and IP setting. The focus on these three categories is consistent with several other papers (15-17). ED, IP, and PCP visits were defined as asthma visits if the primary diagnosis code was asthma. From this point on, we denote asthma-related visits as including ED, IP, and PCP visits unless otherwise stated. We define PCP visits with a place of service code as an “office” or “outpatient hospital.”

The dependent variables consisted of the probability of ED, PCP, and IP visit, utilization expenditures per person per year, and asthma medication expenditures per person per year. We denote utilization expenditures as the combined expenditure for ED, PCP, and IP visits.

We calculate medication costs from the expenditures for controllers and quick reliever medications. For all interventions except seasonal influenza, we consider the intervention and the post-intervention time intervals as the entire calendar year (2010 and 2011, respectively). We assumed that the flu season occurred in the first and last quarters of the calendar year.

### Model Covariates

Each model consisted of a set of independent variables that we evaluated under three categories (demographic characteristics, utilization, and medication adherence). We classified a demographic group as urban, suburban, or rural location. We used two age groups: 0 to 8 years old or 9 to 17 years old. We defined the asthma medication ratio (AMR) (18) for an individual as the ratio of controller medications to total asthma medications. We calculated the use of appropriate medications for people with asthma (ASM) (18) as “1” for children who filled at least one prescription in a calendar year for an asthma controller medication and “0” otherwise. We use medication management for people with asthma (MMA) as another measure that assesses the patients who remained with controller medications at least 50% of their treatment time (18). We used four utilization variables, the number of IP, PCP, ED visits, and non-asthma-related ED visits each year as independent variables.

We analyzed four separate difference-in-difference regression models for each of the dependent variables to estimate the effects of nebulizers, spacers, and AS-ME. We use dependent variables of ED utilization indicator, IP utilization indicator, utilization expenditures, and asthma medication expenditures per person per year. We estimated the effect of influenza vaccines on the PCP utilization indicator, utilization expenditures, and medication expenditure outcome variables. We used asthma medication expenditures in the analysis of NY, and because of insufficient data in MI, we excluded medication expenditures from those models.

We added covariates to each regression model to control for other factors that were not affected by the intervention (19). We included medication adherence, demographic characteristics, and intervention variables in all of the regression models, regardless of the dependent variable. We incorporated utilization variables in the models if they were not related directly to the outcome variable. We dropped non-significant terms from the regressions through a backward stepwise method.

### Regression Methods

We used difference-in-difference (DiD) regression to estimate the intervention’s effect on the probability of asthma-related healthcare utilization and asthma medication and utilization expenditures. We provide parallel trend lines, which show the changes in outcome variables from 2010 to 2011, in Figures 1 and 2 for AS-ME (please refer to Supplementary Figures S1 and S6 for other interventions). For intervention groups, we assessed the average change in outcome measures from pre-intervention/intervention (2010) to post-intervention (2011) periods by comparing this with the average change in the control group over the same time horizon. [Figures 1 and 2 near here]

**Figure 1.**
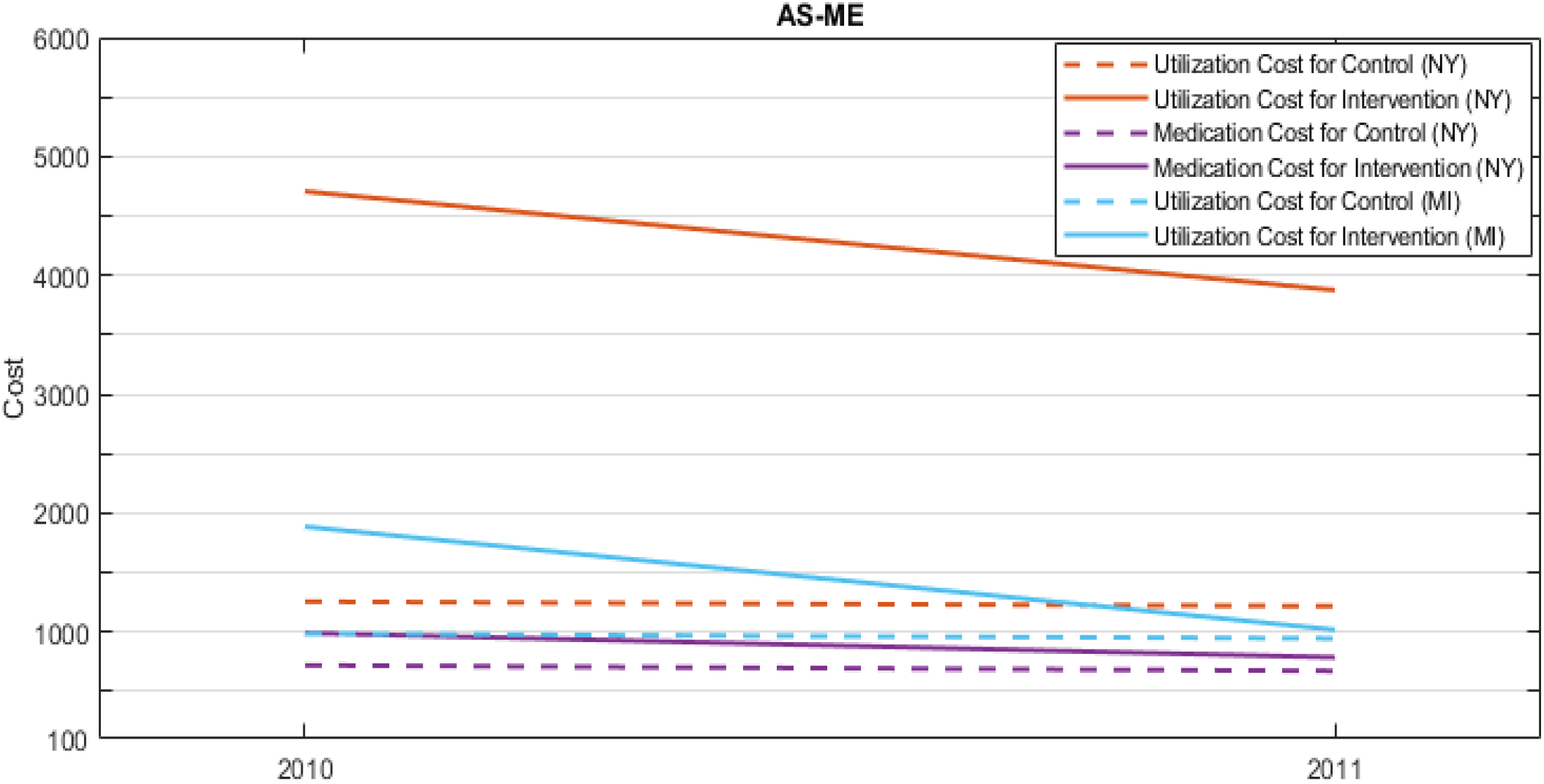
Cost trendlines for AS-ME. Abbreviations: AS-ME - Asthma self-management education

**Figure 2.**
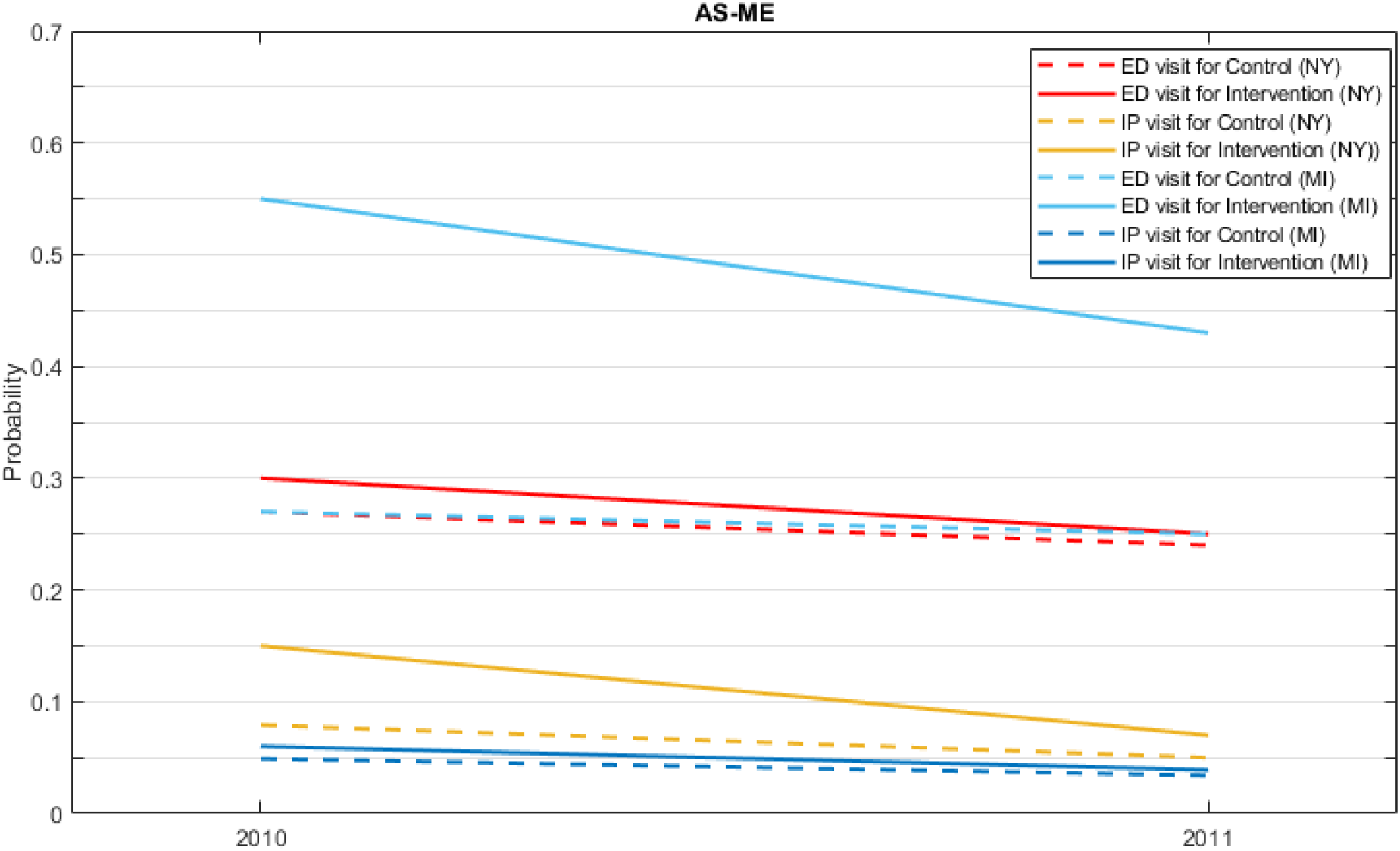
Probability of utilization trendlines for AS-ME. Abbreviations: AS-ME – Asthma self-management education, ED – Emergency department, IP – Inpatient (hospitalization), PCP – Primary care physician

DiD regression required three specific dummy variables, which were related to time and population. We defined a binary intervention variable for each intervention model, which was “1” for the intervention group and “0” for the control group. The year indicator was “1” for 2011 and “0” for 2010. The interaction term represented the interaction between the intervention and the year covariates. The interaction term was defined as the DiD estimator (*ρ*) and predicted with the following equation:

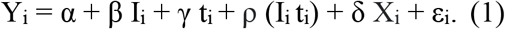

In the equation, the subscript *i* represents the patient; I and t, respectively, represent intervention and time indicators; α is the constant term and represents the intercept; β corresponds to the intervention group effect; γ is time trend for intervention and control groups; *ρ* is the estimator of the intervention effect; *δ* is the effect of time-varying covariates; and ε is the error term. The DiD estimator was calculated from:

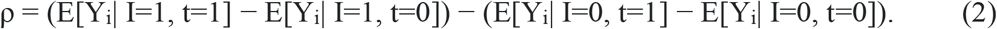

The calculation was obtained by subtracting the change in outcomes for the control group from the change in outcomes for the intervention group over time (20). We used a linear probability model with the untransformed binary outcome variable, and we used an ordinary least squares regression model to estimate log-transformed (natural log) continuous response variables. In order to deal with the log transformation of zero cost values, we added a constant amount of $1 to annual utilization and medication costs before the transformation.

The coefficient of the interaction term represents the effect of an intervention on the outcome variable. Linear probability models can be interpreted directly as the probability of ED, IP, or PCP utilization changes of the intervention population from pre- to post-intervention, over the control group changes. When the interaction term increases from 0 to 1 and all the other variables remained constant, the (DiD estimator × 100) shows the percentage change to the probability of having at least one visit to the specified department, which was indicated as the response variable. In the log-transformed response variable model, if the interaction term increased from 0 to 1 and all the other variables remain constant (e^DiD estimator^ − 1) × 100 (for more information, see (21)), this indicates the percentage of expenditure changes. Negative terms indicate a reduction in expenditures or probability, and a positive sign indicates the opposite. The percentage changes represent the reduction from 2010 to 2011 in the intervention population over the changes in the control population.

## Results

We summarize descriptive results in Tables 1 and 2 for the intervention and control populations. Patients that received interventions were more likely to be younger children, more likely to live in urban or suburban areas, and are likely to have received some other preventive intervention.

**Table 1.**
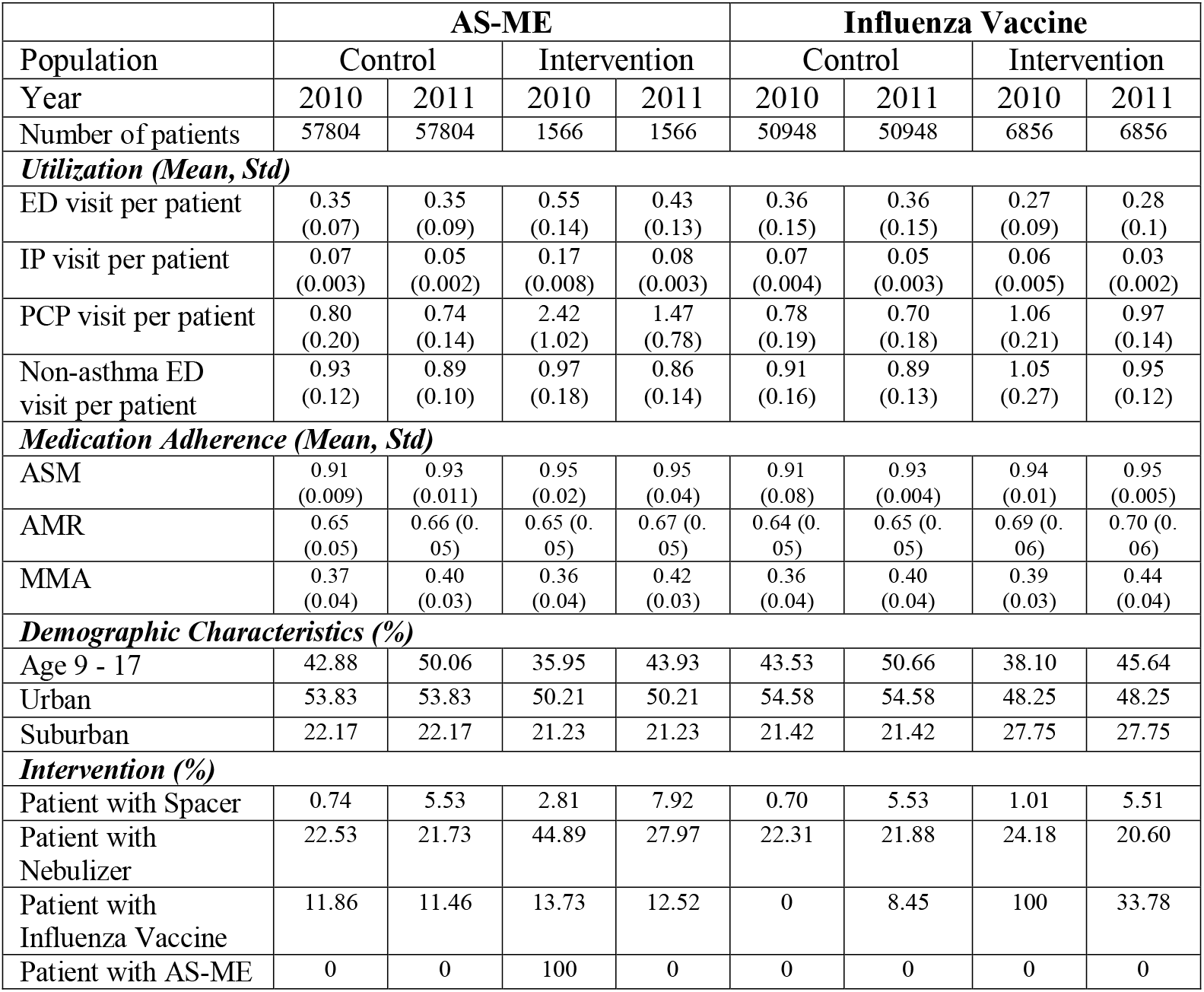
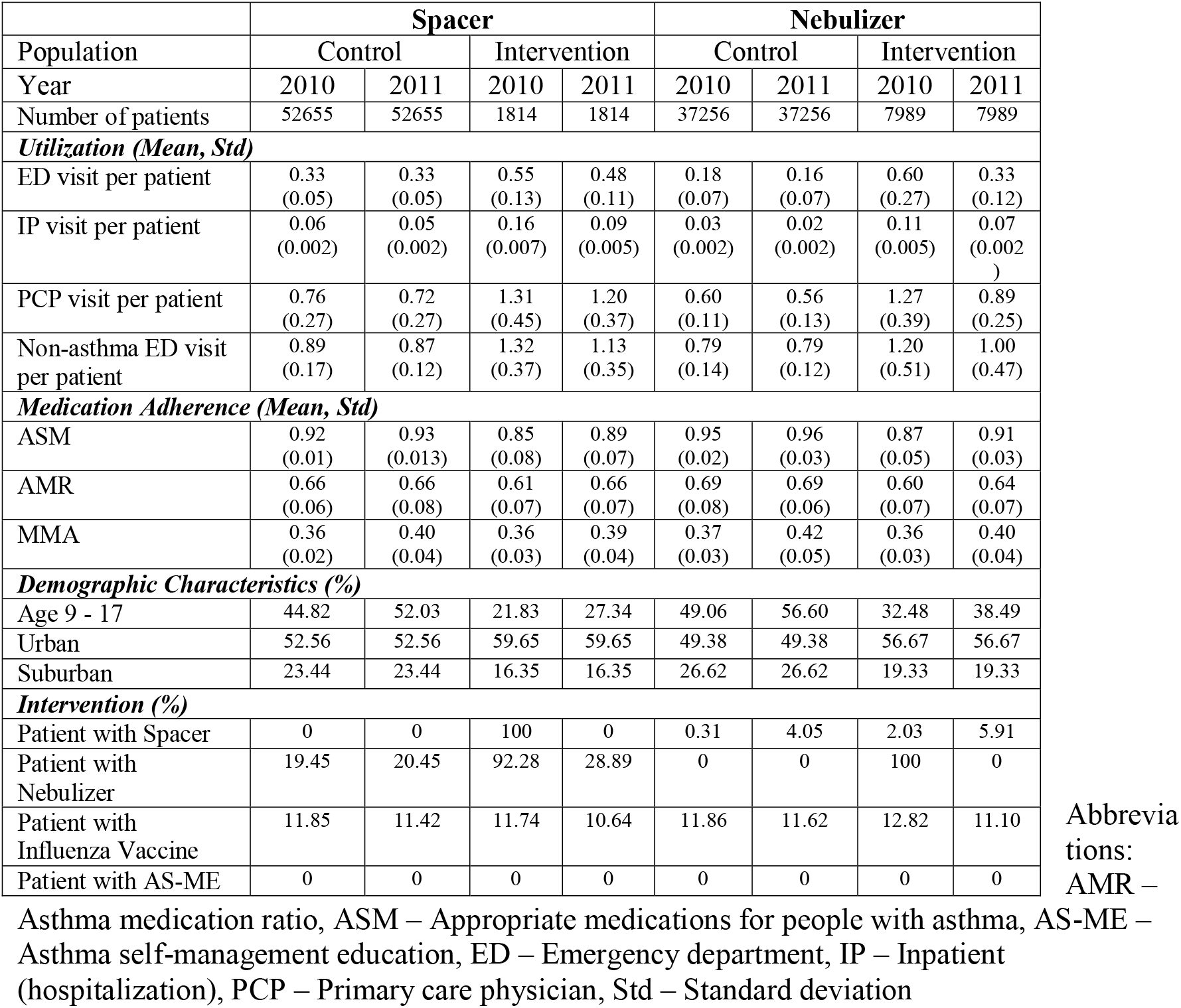
Variable comparison for control and intervention (New York).

**Table 2.** Variable comparison for control and intervention populations (Michigan).

|  | **AS-ME** |  |  |  | **Influenza Vaccine** |  |  |  |
| --- | --- | --- | --- | --- | --- | --- | --- | --- |
| Population | Control |  | Intervention |  | Control |  | Intervention |  |
| Year | 2010 | 2011 | 2010 | 2011 | 2010 | 2011 | 2010 | 2011 |
| Number of patients | 13643 | 13643 | 475 | 475 | 9517 | 9517 | 4126 | 4126 |
| **Utilization (Mean, Std)** |  |  |  |  |  |  |  |  |
| ED visit per patient | 0.36<br>(0.07) | 0.32<br>(0.06) | 0.82<br>(0.23) | 0.41<br>(0.17) | 0.40<br>(0.13) | 0.35<br>(0.11) | 0.28<br>(0.1) | 0.25<br>(0.09) |
| IP visit per patient | 0.05<br>(0.003) | 0.03<br>(0.003) | 0.08<br>(0.004) | 0.04<br>(0.002) | 0.05<br>(0.003) | 0.04<br>(0.003) | 0.04<br>(0.003) | 0.02<br>(0.003) |
| PCP visit per patient | 1.61<br>(0.40) | 1.30<br>(0.28) | 2.21<br>(1.02) | 1.35<br>(0.52) | 1.47<br>(0.24) | 1.22<br>(0.19) | 1.93<br>(0.19) | 1.49<br>(0.38) |
| Non-asthma ED visit per patient | 1.00<br>(0.15) | 0.86<br>(0.10) | 1.26<br>(0.22) | 1.34<br>(0.14) | 0.98<br>(0.12) | 0.84<br>(0.11) | 1.05<br>(0.32) | 0.90<br>(0.31) |
| **Medication Adherence (Mean, Std)** |  |  |  |  |  |  |  |  |
| ASM | 0.28<br>(0.04) | 0.91<br>(0.05) | 0.36<br>(0.09) | 0.91<br>(0.04) | 0.27<br>(0.05) | 0.90<br>(0.02) | 0.30<br>(0.07) | 0.94<br>(0.01) |
| AMR | 0.21<br>(0.01) | 0.63<br>(0.09) | 0.27<br>(0.02) | 0.59<br>(0.1) | 0.20<br>(0.01) | 0.61<br>(0.12) | 0.22<br>(0.01) | 0.68<br>(0.15) |
| MMA | 0.11<br>(0.007) | 0.34<br>(0.05) | 0.16<br>(0.006) | 0.45<br>(0.05) | 0.11<br>(0.004) | 0.33<br>(0.05) | 0.13<br>(0.004) | 0.35<br>(0.06) |
| **Demographic Characteristics (%)** |  |  |  |  |  |  |  |  |
| Age 9 - 17 | 49.1 | 58.5 | 45.5 | 54.9 | 51.5 | 60.5 | 43.5 | 53.8 |
| Urban | 43.0 | 43.0 | 39.6 | 39.6 | 45.4 | 45.4 | 37.7 | 37.7 |
| Suburban | 45.7 | 45.7 | 48.6 | 48.6 | 43.7 | 43.7 | 50.4 | 50.4 |
| **Intervention (%)** |  |  |  |  |  |  |  |  |
| Patient with Spacer | 16.9 | 9.8 | 24.4 | 8.6 | 14.9 | 9.1 | 21.4 | 11.6 |
| Patient with Nebulizer | 43.2 | 34.9 | 59.8 | 40.2 | 43.4 | 35.5 | 42.7 | 33.5 |
| Patient with Influenza Vaccine | 30.2 | 25.5 | 29.1 | 25.5 | 0 | 16.6 | 100 | 46.1 |
| Patient with AS-ME | 0 | 0 | 100 | 0 | 0 | 0 | 0 | 0 |

Table 2. Variable comparison for control and intervention populations (Michigan) (continued).
|  | **Spacer** |  |  |  | **Nebulizer** |  |  |  |
| --- | --- | --- | --- | --- | --- | --- | --- | --- |
| Population | Control |  | Intervention |  | Control |  | Intervention |  |
| Year | 2010 | 2011 | 2010 | 2011 | 2010 | 2011 | 2010 | 2011 |
| Number of patients | 10426 | 10426 | 1875 | 1875 | 5980 | 5980 | 2903 | 2903 |
| **Utilization (Mean, Std)** |  |  |  |  |  |  |  |  |
| ED visit per patient | 0.36<br>(0.05) | 0.31<br>(0.05) | 0.35<br>(0.05) | 0.28<br>(0.03) | 0.62<br>(0.28) | 0.55<br>(0.11) | 0.50<br>(0.09) | 0.12<br>(0.04) |
| IP visit per patient | 0.04<br>(0.002) | 0.03<br>(0.002) | 0.05<br>(0.002) | 0.03<br>(0.002) | 0.07<br>(0.002) | 0.05<br>(0.002) | 0.05<br>(0.002) | 0.02<br>(0.002) |
| PCP visit per patient | 1.56<br>(0.41) | 1.24<br>(0.38) | 1.92<br>(0.4) | 1.26<br>(0.22) | 2.72<br>(1.02) | 2.16<br>(0.81) | 1.92<br>(0.52) | 1.12<br>(0.37) |
| Non-asthma ED visit per patient | 0.94<br>(0.17) | 0.81<br>(0.12) | 1.11<br>(0.29) | 0.42<br>(0.32) | 0.62<br>(0.14) | 0.52<br>(0.12) | 1.13<br>(0.38) | 1.23<br>(0.31) |
| **Medication Adherence (Mean, Std)** |  |  |  |  |  |  |  |  |
| ASM | 0.27<br>(0.09) | 0.90<br>(0.07) | 0.38<br>(0.08) | 0.95<br>(0.03) | 0.30<br>(0.1) | 0.99<br>(0.008) | 0.28<br>(0.07) | 0.95<br>(0.03) |
| AMR | 0.20<br>(0.01) | 0.62<br>(0.07) | 0.27<br>(0.01) | 0.67<br>(0.08) | 0.23<br>(0.01) | 0.69<br>(0.09) | 0.20<br>(0.01) | 0.67<br>(0.1) |
| MMA | 0.11<br>(0.009) | 0.34<br>(0.04) | 0.17<br>(0.006) | 0.34<br>(0.09) | 0.12<br>(0.006) | 0.36<br>(0.01) | 0.12<br>(0.006) | 0.35<br>(0.01) |
| **Demographic Characteristics (%)** |  |  |  |  |  |  |  |  |
| Age 9 - 17 | 54.5 | 63.7 | 31.9 | 42.5 | 58.8 | 68.5 | 43.6 | 53.1 |
| Urban | 44.3 | 44.3 | 38.1 | 38.1 | 35.0 | 35.0 | 42.5 | 42.5 |
| Suburban | 44.2 | 44.2 | 50.6 | 50.6 | 51.0 | 51.0 | 46.2 | 46.2 |
| **Intervention (%)** |  |  |  |  |  |  |  |  |
| Patient with Spacer | 0 | 0 | 100 | 0 | 14.7 | 7.6 | 20.5 | 9.6 |
| Patient with Nebulizer | 41.1 | 33.6 | 49.2 | 34.5 | 0 | 0 | 100 | 0 |
| Patient with Influenza Vaccine | 28.2 | 24.5 | 37.5 | 28.5 | 30.7 | 25.7 | 31.3 | 25.9 |
| Patient with AS-ME | 0 | 0 | 0 | 0 | 0 | 0 | 0 | 0 |

Abbreviation
s: AMR – Asthma medication ratio, ASM – Appropriate medications for people with asthma, AS-ME – Asthma self-management education, ED – Emergency department, IP – Inpatient (hospitalization), PCP – Primary care physician, Std – Standard deviation

For patients who received any intervention, their 2010 utilization cost was 1.37 times higher than those who received none. In comparison to the control, their probability of ED, IP, and PCP utilization was 41% higher, 122% higher, and 55% lower in 2010.

For each intervention and each outcome variable, we estimated the intervention effect for patients who received an intervention in 2010. For all interventions in both states, we find that the utilization cost decreased after the intervention, which is illustrated in Figures 3 and 4. These figures convert the DiD estimator to the % of change values provided from Tables S1 and S2 (see Supplementary file). We discuss results specific to each intervention below.

**Figure 3.**
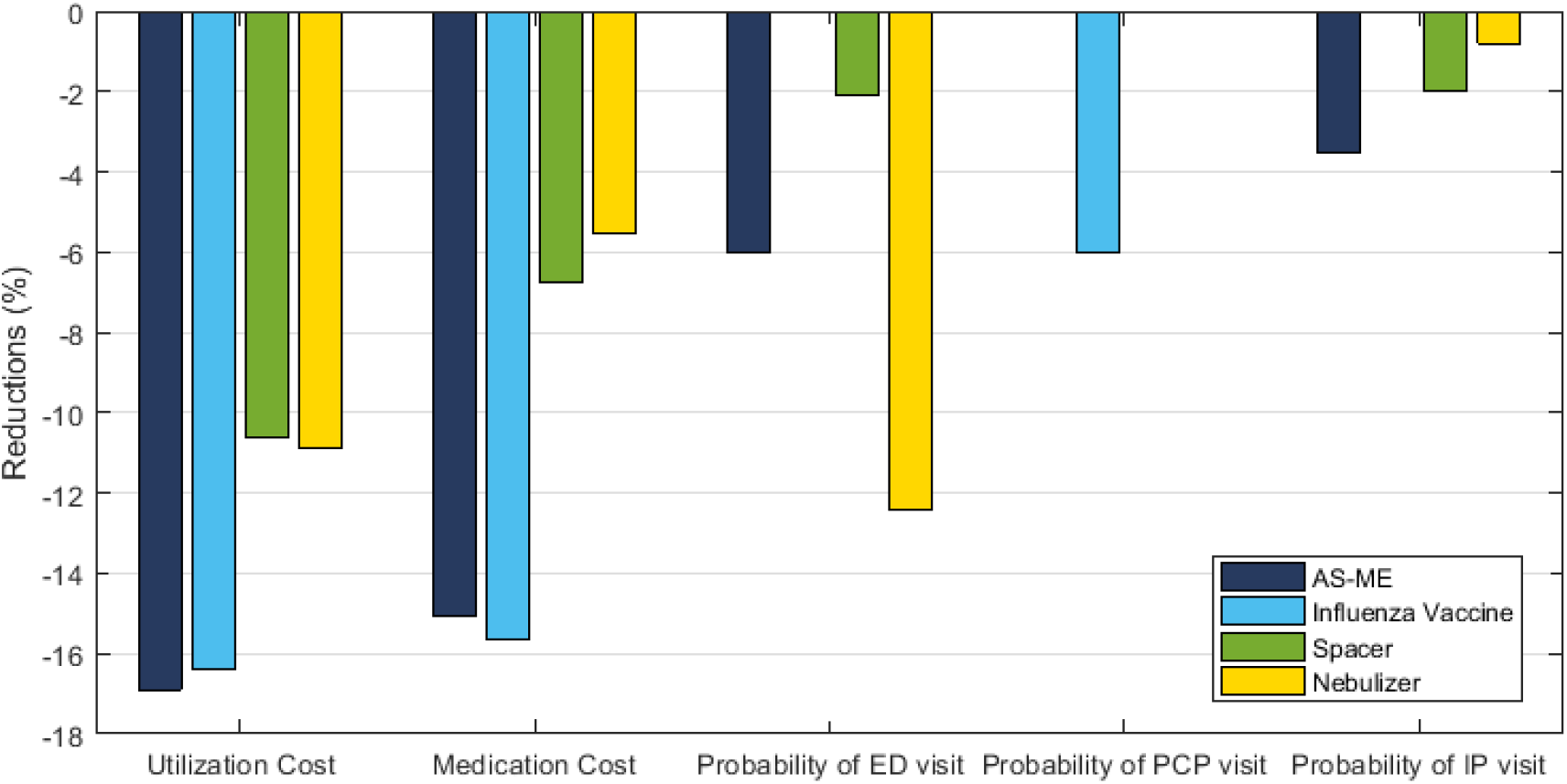
Summary results for intervention effects (New York). Notes: Calculations performed with using Supplementary Tables S1 and S2. Utilizations are transformed to % changes by DiD estimator × 100 and costs are transformed with using e^DiD estimator^ − 1 × 100. Abbreviations: AS-ME – Asthma self-management education, ED – Emergency department, IP – Inpatient (hospitalization), PCP – Primary care physician

**Figure 4.**
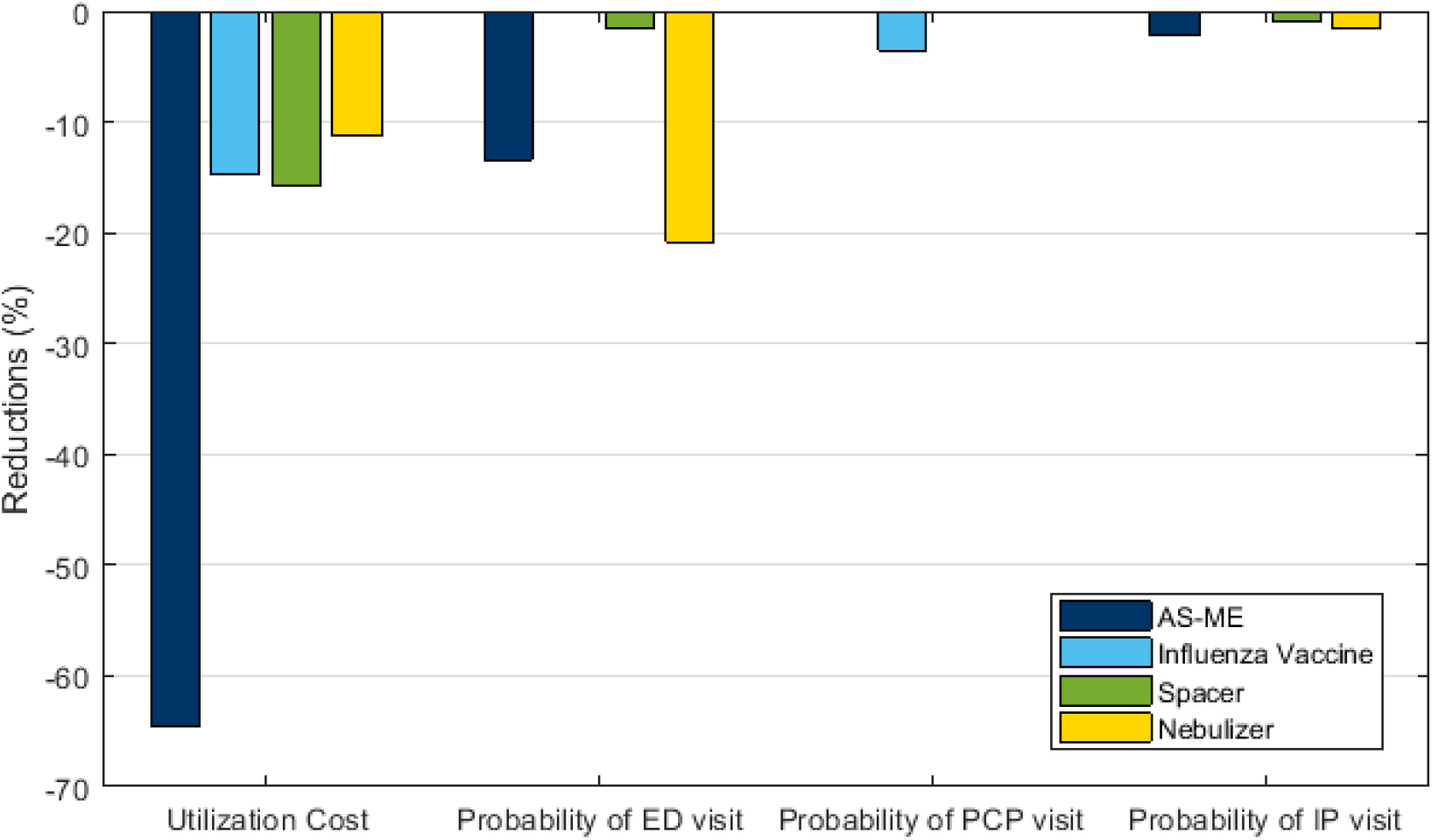
Summary results for intervention effects (Michigan). Notes: Calculations performed with using Supplementary Tables S1 and S2. Utilizations are transformed to % changes by DiD estimator × 100 and costs are transformed with using e^DiD estimator^ − 1 × 100. Abbreviations: AS-ME – Asthma self-management education, ED – Emergency department, IP – Inpatient (hospitalization), PCP – Primary care physician

### AS-ME

In the data, patients who were in the intervention group for AS-ME showed a significant reduction in ED utilization from 2010 to 2011. For example, the intervention group made 0.82 ED visits in MI and 0.55 in NY on average in 2010, while in 2011, their average number of ED visits decreased to 0.41 in MI and 0.43 in NY. On the other hand, the AS-ME control population had similar average ED utilization per patient in both 2010 and 2011.

Regression results show that AS-ME decreased the probability of ED utilization by 13.4% in MI and 6% in NY and IP utilization by 2.2% in MI and 3.5% in NY. Figures 3 and 4 summarize the results, showing that AS-ME decreased the utilization cost by 16.9% (i.e., =(e^-0.185^ − 1) × 100, using coefficient −0.185 from Table S2) for NY and 64.6% for MI patients. The NY data shows that AS-ME reduced asthma medication expenditures (controller plus reliever expenditures) by 15.04%. [Figures 3 and 4 near here]

### Influenza Vaccine

From descriptive data, patients in the intervention group had 26.4% and 23.8% (0.78 control vs. 1.06 intervention and 1.47 control vs. 1.93 intervention) higher PCP utilization than the control populations, respectively, in NY and MI in 2010. Both the control and intervention populations had decreased mean PCP utilization in 2011. On the other hand, the intervention for influenza vaccine had decrease average IP utilization per patient in NY and MI by 50% (0.06 to 0.03 in NY and 0.04 to 0.02 in MI) from 2010 to 2011.

Regression results estimate that patients who received the influenza vaccine in 2010 reduced their utilization expenditures by 16.4% in NY and 14.8% in MI. The vaccination was associated with a 6% (NY) and 3.5% (MI) lower probability of a PCP visit. Furthermore, the influenza vaccine decreased asthma medication expenditures by 15.6% in NY (Figures 1 and 2).

## Nebulizer

Descriptive data from Tables 1 and 2 show that the mean ED utilization reduced 45% in MI (76% in NY) in the intervention population, which is 0.50 vs. 0.12 (0.60 vs. 0.33), respectively, in 2010 and 2011. The nebulizer control population had 11% (0.18 vs. 0.16 in NY and 0.62 vs. 0.55 in MI) difference in mean ED utilization compared to the intervention population in both states.

From the regression models, the 2011 reduction of the predicted ED utilization probability was 20.8% in MI and 12.4% in NY with the nebulizer intervention. In addition, nebulizers reduced the likelihood of IP utilization by 1.6% and 0.8%, respectively, in MI and NY. [Figures 1 and 2 near here] Nebulizers decreased the utilization cost around 11% in both states and medication cost 6% in NY.

### Spacer

In both states, the intervention population for spacers reduced their mean ED and IP utilization from 2010 to 2011 more than the reduction of the control group. The mean ED utilization was reduced 20% (12.7%) for the intervention population (0.35 vs. 0.28 in MI and 0.55 vs. 0.48 in NY) in MI (NY) versus 13.9% (0%) in the control population (0.36 vs. 0.31 in MI and 0.33 vs. 0.33 in NY). Similarly, the reduction of mean IP utilization was 43.8% (0.16 vs. 0.09) and 40% (0.05 vs. 0.03) for the intervention population, whereas, it was 16.7% (0.06 vs. 0.05) and 25% (0.04 vs. 0.03) for the control population, respectively, in NY and MI (Tables 1 and 2).

Having a spacer decreased medication and utilization expenditures. The reduction of ED and IP utilization probability was around 1% in MI and 2% in NY. A spacer decreased the utilization cost by 10.6% for NY and 15.7% for MI patients.

## Discussion

All the interventions reduced the annual utilization and asthma medication expenditures per intervention patient. AS-ME, nebulizer, and spacer interventions reduced the probability of ED and IP utilization. Influenza vaccines decreased the likelihood of PCP utilization. In short, all of the interventions had positive impacts on more than one measure of outcomes or cost. This is true even when controlling for other interventions that a patient had, which commonly occurred and when controlling for regression to the mean through the difference-in-difference approach.

The percentages of expenditure reductions were consistent for the two states for influenza vaccine (15% in MI and 16% in NY) and nebulizer (11% in MI and NY) models. The spacer intervention population showed similar reductions for two states (16% in MI and 11% in NY). However, AS-ME results showed a 65% expenditure reduction in MI and a 17% reduction in NY.

A difference between programs is one potential explanation. AS-ME implementation processes differ from many perspectives; however, we can observe only some of them from claims data (e.g., place of service, provider). Specifically, 77% of AS-ME claims in NY were in a Physician’s office, whereas most of them (65%) in MI took place in outpatient hospitals. In NY, pediatricians (70%) were commonly responsible for AS-ME while in MI, the primary care physicians (60%) were. Additionally, intervention populations in NY and MI were different based on their average utilizations in 2010. In these states, AS-ME may have been given to different patient groups based on the severity of asthma. Some of these differences may have been due to differences in procedure coding systems, state regulations, or rules that determined the target populations for the interventions.

In most cases, the reductions in the probability of healthcare utilization are consistent from state to state. The results from nebulizer utilization models showed that with the nebulizer intervention, NY patients were five times more likely to decrease IP utilization and 1.7 times less likely to reduce ED utilization compared to the MI intervention patients. This suggests that there may be a shifting of healthcare resources, e.g., a proportion of IP visits are handled in the ED, and ED visits are reduced with more PCP visits.

Our results on AS-ME are consistent with previous studies. AS-ME can reduce ED and IP utilization (22-25), decrease the cost of utilization, improve medication adherence (ASM, AMR, MMA in Tables 1 and 2), and reduce asthma medication costs, which can be caused by excessive use of relievers. Lower AMRs indicate the possibility of higher consumption of relievers. AS-ME improves controller adherence since MMA increases with AS-ME patients more than control patients. Whereas, the number of claims drops for short-acting inhaled beta-agonists (SABA), which is associated with increased AMR.

The results show that the effect of nebulizer and spacer in the treatment of asthma was similar to other studies (26,27). There was limited evidence for the impact either way of the influenza vaccine on asthma exacerbations in pediatric asthma patients (28). Further, some studies showed no effect from the influenza vaccine on health outcomes for children with asthma (29-31). However, a flu infection can trigger asthma attacks and a need for quick relievers (12). Patients who received an influenza vaccine may decrease their medication costs due to an influenza case averted or shortened by the vaccine.

The raw data in Tables 1 and 2 showed that the percentage reduction in “average” IP visits is more significant than the percentage reduction in “average” ED visits for some interventions (nebulizer for both states and AS-ME in NY). However, Figures 3 and 4 showed that the percentage probability of a reduction in IP utilization is lower than the percentage probability of a reduction in ED visits. There may be a couple of reasons for this. An important one is that the regressions that we use in this study account for other covariates. As mentioned earlier, there may also be additional differences in implementation in the specific states or for a given intervention.

Influenza vaccines ($12.45-$32.47 (32)) and spacers ($4-$30 (33)) are not costly investments. While AS-ME interventions are more costly, the AS-ME intervention population had the highest utilization expenditure reduction compared to the other intervention populations.

We did not use matching methods to create a control group because of two reasons. First, even if control and intervention populations do not have similar pre-intervention means, DiD models can generate unbiased estimates (34). Secondly, matching the control and intervention population can lead to a biased assessment because regression to the mean occurred when the population was selected for a higher/lower-than-average in the first measurement (2010) (35). The matched population will tend to regress to the population mean in the second year (2011).

The study has limitations. The data analysis started in 2016 with the latest available MAX data from 2010 and 2011. The results of models depended on the available data. We were only able to control for certain factors, and it is possible that unobserved factors could affect the expenditures or utilizations. However, DiD is a useful tool because it eliminates the need to control all confounding variables (19). In this study, we consider both population differences (demographics, medication adherence, utilization) and various interventions within the same model. The intervention and control patients differed from each other based on the utilization of services in 2011. These differences could have originated from the intervention and other factors (any other asthma control efforts not seen in the data). We did not evaluate the effects of intervention combinations (e.g., AS-ME + Spacer) due to the small sample sizes of the subpopulations. Throughout the analysis, we did not specify the type of medication (controller, reliever) obtained with the spacer and nebulizer. The HEDIS measure we use excludes systematic corticosteroids, which could lead to a misestimation of medication costs.

We assume that the patients who have a claim for intervention indeed receive the intervention. On the other hand, patients defined for the control group never received that intervention, while some may have received it from other sources (e.g., spacer, influenza vaccine). We are accepting that uncertainty can exist, and we acknowledge that statistical analysis does not always indicate an essential relationship. The evaluation of observed results from clinical trials could give more precise estimates about effects and associations (36,37).

## Conclusions

Previous studies (17,22,25-29,31), literature reviews or meta-analysis (23,24,30), showed the impact of asthma interventions on a population by looking at them individually, which can lead to under- or over-estimating the set of effects taken in total. In this study, we consider both population differences (demographics, medication adherence, utilization) and various interventions within the same model in order to develop a better estimate of the overall effect of multiple interventions.

This analysis provides evidence to policymakers about the benefits of the interventions of influenza vaccines, spacers, AS-ME, and nebulizers on health outcomes of pediatric asthma patients. Influenza vaccines were shown to be the most effective intervention on medication expenditures. AS-ME programs showed the highest utilization expenditure reductions overall. AS-ME also improved medication adherence and reduced asthma medication costs caused by the utilization of rescue medications. The spacer and nebulizer decreased utilization expenditure, which was caused by the reduction in the probability of ED and IP utilization. Although the percentage of patients that benefited from the interventions were low, promoting these interventions in other states or health systems could decrease the utilization cost and the frequency of healthcare utilization by the sickest patients while improving medication compliance of patients.

## Data Availability

Medicaid claims data were available at Georgia Tech
through support by the Institute for People and Technology, Stewart School of Industrial and Systems Engineering, and Childrens Healthcare of Atlanta.

## Notes

### Competing Interest Statement

The authors have declared no competing interest.

### Funding Statement

P. Griffin was supported by Regenstreif Center. P.Keskinocak received partial support from William W. George chair. J.Swann received partial support from A. Doug Allison chair.

### Author Declarations

The IRB that ruled on the ethics of our study was the Georgia Tech Institute Review Board (IRB) for Georgia Tech Research Corporation. Ethical approval was given by IRB for the research.

